# Enhanced precision of tensor electrocardiography through increased cumulative distribution function resolution: Validation in healthy individuals

**DOI:** 10.64898/2026.05.31.26354561

**Authors:** Yayoi Tetsuo Tsukada, Hiroaki Hirayama, Kenji Yodogawa, Hiroshige Murata, Yu-ki Iwasaki, Takeo Fujino, Akihiro Shiozawa, Shingo Tsukada

## Abstract

Deep-learning ECG analysis is advancing rapidly but lacks stable, physiologically interpretable indicators to anchor explainable artificial intelligence (AI). Tensor cardiography (TCG) models electrocardiographic (ECG) waveforms as differences between pairs of cumulative distribution functions (CDFs), representing collective myocardial action potential transitions. However, the original 4-CDF model has limitations in fitting P waves and complex QRST patterns. This study aimed to evaluate whether increasing the number of CDFs from 4 to 10 improves TCG fitting accuracy and to characterize normative distributions of 10-CDF parameters in healthy individuals.

Participants were recruited through occupational health screening at Tobu Railway Co., Ltd. (n = 415) and from the Nippon Medical School Hospital ECG database (n = 29). Standard 12-lead ECGs from 444 healthy participants, including 345 men and 99 women with a mean age of 46.9 years, were analyzed using TCG software. Reconstruction accuracy was assessed using RMSE, paired t-tests, and Cohen’s d.

The 10-CDF model achieved significantly lower RMSE values across all leads than the 4-CDF model, with all p values < 0.0001 and very large effect sizes. In representative leads, RMSEs for the 4-CDF versus 10-CDF models were 0.0256 versus 0.0061 in lead II, 0.0230 versus 0.0063 in lead V1, and 0.0265 versus 0.0062 in lead V5. The coefficient of determination improved from a median of 0.952 with the 4-CDF model to 0.997 with the 10-CDF model in lead II. Parameter dispersion was reduced, suggesting improved estimation stability. Two new parameters, T_mean_diff and RT_mean_duration, were derivable from the expanded model; RT_mean_duration showed significant correlations with age and body surface area.

In conclusion, increasing the CDF resolution from 4 to 10 significantly enhanced ECG waveform reconstruction accuracy and parameter stability. These findings provide normative distributions of 10-CDF TCG parameters and may support future explainable AI-based ECG analysis.

**Author summary:** Deep-learning approaches have rapidly advanced electrocardiogram (ECG) analysis, but many models function as black boxes with indicators that are difficult for clinicians to interpret. There is a growing need for stable, physiologically meaningful ECG measurements to make artificial intelligence (AI) more explainable. Tensor cardiography (TCG) addresses this need by representing ECG waveforms using cumulative distribution functions (CDFs) based on myocardial electrical activity models, providing interpretable parameters. In this study, we increased the number of CDFs from four to ten to extract more precise quantitative information from ECGs of 444 healthy individuals. The 10-CDF model reconstructed ECG waveforms with substantially greater accuracy than the original 4-CDF model, reducing fitting errors by approximately 75%. We also characterized normative reference distributions for the expanded model and identified two new parameters that capture details of cardiac electrical activity not available in the original 4-CDF model. These findings suggest that the expanded TCG model provides a stable, interpretable framework for future AI-assisted and wearable ECG analysis. By linking waveforms to physiological parameters, TCG may help support earlier and more transparent detection of heart disease.

## Introduction

Electrocardiograms (ECG) have been indispensable diagnostic tools in cardiovascular medicine for over a century, providing critical information regarding cardiac rhythm, conduction, and myocardial function [1]. Despite its ubiquity, quantitative analysis of ECG waveforms remains challenging. Conventional ECG interpretation relies primarily on measuring discrete intervals and amplitudes supplemented by morphological pattern recognition [2]. Although automated diagnostic algorithms have improved interpretation efficiency, they largely depend on rule-based criteria that do not fully exploit the rich information encoded in continuous waveform morphology [3].

In this context, mathematical modeling approaches have been used to address these limitations. For instance, transform-based methods, including discrete cosine transform (DCT) and wavelet transform-based analyses, have been utilized for the detection of life-threatening arrhythmias and risk stratification in patients at risk of ventricular arrhythmias [4,5]. Discrete wavelet transforms (DWTs) have also been widely used in ECG signal compression and reconstruction [6,7]. However, these engineering-oriented approaches lack high predictive accuracy and produce coefficients that lack direct physiological interpretability, limiting their clinical utility beyond data storage and transmission.

More recently, deep-learning approaches — typified by one-dimensional convolutional architectures such as ResNet and Inception trained on PTB-XL — have achieved high benchmark performance in automated ECG classification [8], with parallel developments across time-, frequency-, decomposition-, and tensor-based signal representations [9]. However, these models typically demand substantial computational resources and offer limited explainability: while explainable-by-design strategies such as variational auto-encoders have been proposed [10], post-hoc saliency methods can be unstable and only modestly informative to clinicians [11]. Recent work has shown that compact, physiologically grounded features can approach deep-learning accuracy while remaining interpretable [12], a property that is increasingly required as wearable and consumer-grade ECG devices come into routine clinical use [13]. There is therefore a growing call for stable, interpretable ECG indicators that can serve as anchors for explainable artificial intelligence (AI).

Tensor cardiography (TCG), recently introduced by Tsukada et al. [14], offers a fundamentally different approach. TCG models ECG waveforms as the difference between pairs of cumulative distribution functions (CDFs) that represent the collective transitions of myocardial action potentials. The QRS complex is represented by the difference between two CDFs corresponding to the anodic (endocardial) and cathodic (epicardial) components of depolarization, whereas the T wave is similarly decomposed into two CDFs for the repolarization process. Each CDF is characterized by four physiologically interpretable parameters: mean (μ), representing the central timing of the transition; standard deviation (σ), representing temporal dispersion; weight (κ), reflecting amplitude contribution; and baseline level (β) [14]. Consequently, TCG has demonstrated promise in detecting myocardial ischemia and evaluating beat-to-beat repolarization dynamics and an established model using TCG has enabled highly accurate fitting of electrocardiographic waveforms [14].

Nevertheless, the original 4-CDF TCG model has inherent limitations. Even in healthy individuals, with only four CDFs allocated to the QRS and T wave segments, the model has a limited capacity to represent complex waveform morphologies, including the P wave, multiphasic QRS complexes (e.g., RSR patterns in lead V1), notched T waves, and other morphological variants commonly encountered in clinical practice. Increasing the number of CDFs provides additional degrees of freedom for waveform approximation, which may potentially capture all the components of the P-QRS-T waveform. However, normative reference values for higher-resolution CDF models have not yet been established.

Therefore, this study aimed to (1) evaluate whether increasing the number of CDFs from 4 to 10 significantly improves TCG fitting accuracy across the standard 12-lead ECG in a large cohort of healthy individuals; (2) quantify the magnitude of improvement using paired t-tests and effect size measures; (3) establish normative reference values for the 10-CDF TCG parameters, including new parameters uniquely derived from the expanded model; and (4) characterize the relationships of TCG parameters with age and body surface area.

## Results

### Study population characteristics

The study cohort comprised 444 healthy participants (345 men [77.7%] and 99 women [22.3%]) whose demographic characteristics are summarized in Table 1. The cohort had 128, 277, and 39 participants aged 20–39, 40–59, and 60–79 years, respectively.

**Table 1.** Baseline characteristics of the study population (N=444)

| Characteristic | Male (n=345) | Female (n=99) | Total (N=444) |
| --- | --- | --- | --- |
| Age (years), mean | 46.15 | 49.47 | 46.90 |
| Age 20-39, n | 110 | 18 | 128 |
| Age 40-59, n | 207 | 70 | 277 |
| Age 60-79, n | 28 | 11 | 39 |
| Height (cm), mean | 171.1 | 158.6 | 168.8 |
| Weight (kg), mean | 64.7 | 52.6 | 62.4 |
| BMI (kg/m<sup>2</sup>), mean | 22.1 | 20.9 | 21.9 |
| BSA (m<sup>2</sup>), mean | 1.76 | 1.52 | 1.71 |

*BMI, body mass index; BSA, body surface area. Data primarily from the Tobu Railway Co., Ltd. occupational health screening cohort (n=415). Nippon Medical School Hospital subjects (n=29) contributed to TCG analysis.*

### RMSE comparison: 4-CDF vs. 10-CDF model

The 10-CDF TCG model achieved a significantly lower root mean square error (RMSE) than the 4-CDF model across all analyzed leads (all p < 0.0001). The results for the representative leads are listed in Table 2. All three leads had large effect sizes (Cohen’s d=-1.84 ∼ -2.77) for. The representative waveform reconstructions using both models are illustrated in Fig 1.

**Table 2.** RMSE values and paired t-test results: 4-CDF vs. 10-CDF TCG (N=444)

| Lead | 4-CDF RMSE, mean ± SD | 10-CDF RMSE, mean ± SD | t statistic | Cohen's d | p-value |
| --- | --- | --- | --- | --- | --- |
| II | 0.0256 ± 0.0075 | 0.0061 ± 0.0017 | -58.35 | -2.77 | < 0.0001 |
| V1 | 0.0230 ± 0.0089 | 0.0063 ± 0.0023 | -40.44 | -1.92 | < 0.0001 |
| V5 | 0.0265 ± 0.0120 | 0.0062 ± 0.0017 | -38.69 | -1.84 | < 0.0001 |

RMSE values are in normalized units (mV). Cohen's d classification: $|d| > 0.8$ = large effect size. Cohen's d calculated as mean paired difference / SD of paired differences. Analyses performed using paired t-tests ( $df=443$ ) in JMP Pro. CDF, cumulative distribution function; RMSE, root mean square error; SD, standard deviation.

**Fig 1.**
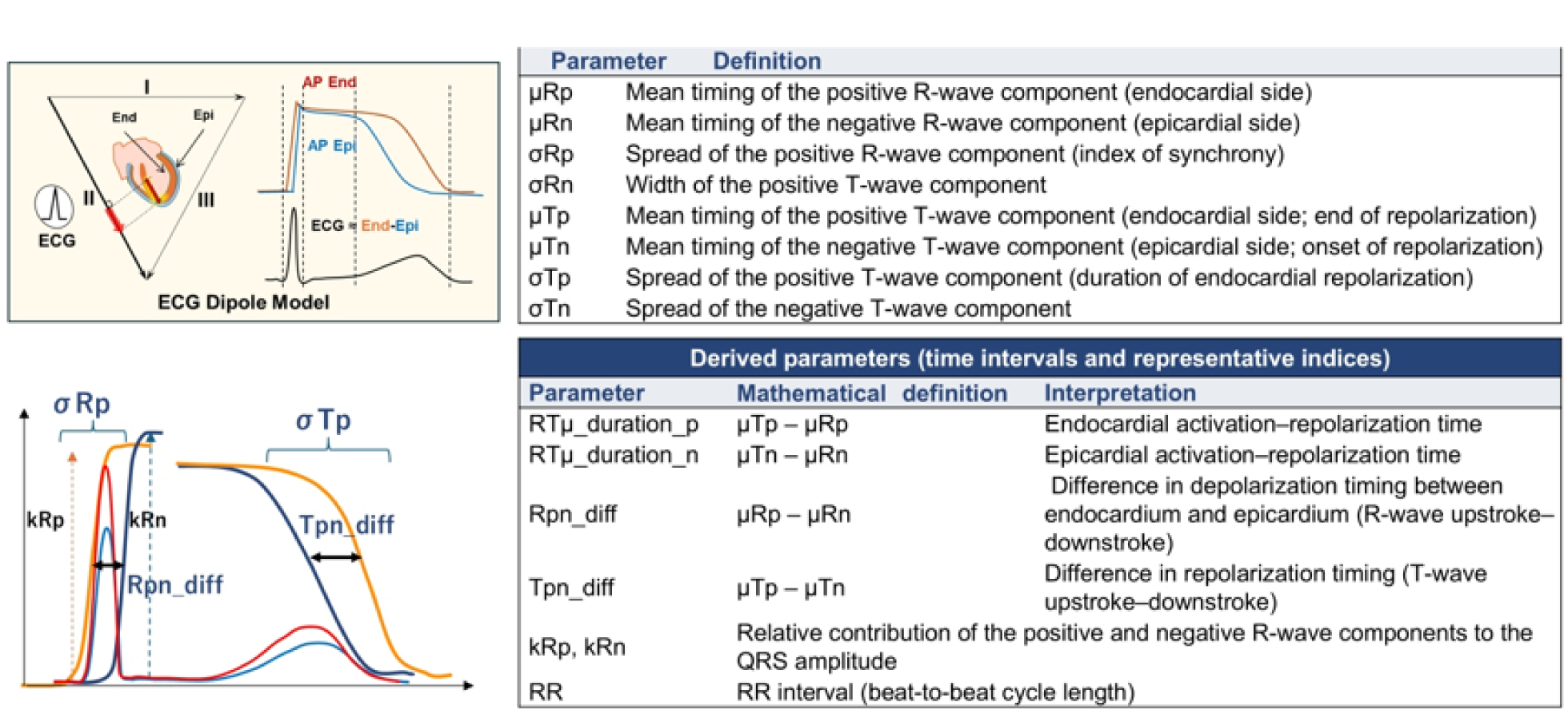
R- and T-wave model parameters and derived indices.

The coefficient of determination (r²) for waveform reconstruction improved dramatically with the 10-CDF model. In lead II, the median r² increased from 0.9516 (4-CDF) to 0.9968 (10-CDF), and in lead V1, it increased from 0.9707 to 0.9980. These results indicate that the 10-CDF model explains > 99.6% of the variance in the ECG waveform compared to 95.2–97.1% for the 4-CDF model.

### Comprehensive parameter comparison: 4-CDF vs. 10-CDF

Table 3 presents a complete parameter comparison between the 4-CDF and 10-CDF models for leads II, V1, and V5. Markedly reduced standard deviations were observed with the 10-CDF model for most parameters (e.g., R_sigma_p SD: 5.50→1.68 in lead II; T_sigma_n SD: 19.95→12.16 in lead II), indicating more stable fitting. The T_sigma_n was significantly lower in the 10-CDF model in leads II and V5 than that in the 4-CDF model (p < 0.0001), suggesting the 4-CDF model systematically overestimates T-wave dispersion. The RT_mean_duration_n was significantly longer in the 10-CDF model in leads II and V5 than that in the 4-CDF model (p < 0.0001). Additionally, the T_mean_diff magnitude was consistently smaller with reduced SD in the 10-CDF model than that in the 4-CDF model, indicating more precise T-wave characterization with the 10-CDF model.

**Table 3.** TCG parameter comparison: 4-CDF vs. 10-CDF (N=444).

| Lead | Parameter | 4-CDF Mean ± | 10-CDF Mean | p-value |
| --- | --- | --- | --- | --- |

|  |  | **SD** | **± SD** |  |
| --- | --- | --- | --- | --- |
| II | R_sigma_p (ms) | 7.09 ± 5.5 | 7.06 ± 1.68 | 0.933 |
|  | R_sigma_n (ms) | 7.64 ± 4.44 | 6.86 ± 2.37 | < .0001 |
|  | T_sigma_p (ms) | 27.56 ± 7.96 | 25.02 ± 4.08 | < .0001 |
|  | T_sigma_n (ms) | 66.59 ± 19.95 | 43.44 ± 12.16 | < .0001 |
|  | RT_mean_duration_p (ms) | 307.58 ± 32.69 | 306.56 ± 26.89 | 0.274 |
|  | RT_mean_duration_n (ms) | 208.51 ± 38.87 | 217.54 ± 24.1 | < .0001 |
|  | QT_mean_duration (ms) | — | 340.6 ± 35.38 | — |
|  | R_mean_diff (ms) | 20.79 ± 5.91 | 20.98 ± 4.26 | 0.454 |
|  | RR (ms) | 917.64 ± 138.06 | 917.79 ± 138.23 | 0.304 |
|  | R_k_p | 0.87 ± 0.37 | 0.91 ± 0.35 | < .0001 |
|  | R_k_n | 0.92 ± 0.35 | 0.87 ± 0.34 | < .0001 |
|  | RMSE | 0.025 ± 0.007 | 0.006 ± 0.001 | < .0001 |
| V1 | R_sigma_p (ms) | 6.01 ± 3.1 | 7.23 ± 2.57 | < .0001 |
|  | R_sigma_n (ms) | 18.87 ± 9.21 | 16.46 ± 6.6 | < .0001 |
|  | T_sigma_p (ms) | 77.86 ± 36.47 | 40.01 ± 19.31 | < .0001 |
|  | T_sigma_n (ms) | 39.78 ± 20.82 | 30.18 ± 9.66 | < .0001 |
|  | RT_mean_duration_p (ms) | 184.24 ± 85.19 | 229.87 ± 56.07 | < .0001 |
|  | RT_mean_duration_n (ms) | 229.25 ± 45.56 | 232.63 ± 28.59 | 0.059 |
|  | QT_mean_duration (ms) | — | 305.61 ± 31.43 | — |
|  | R_mean_diff (ms) | 25.87 ± 10.69 | 32.43 ± 9.46 | < .0001 |
|  | RR (ms) | 917.67 ± 137.91 | 917.86 ± 138.07 | 0.219 |
|  | R_k_p | 1.17 ± 0.49 | 1.1 ± 0.49 | < .0001 |
|  | R_k_n | 1.15 ± 0.48 | 1.07 ± 0.49 | < .0001 |
|  | RMSE | 0.023 ± 0.008 | 0.006 ± 0.002 | < .0001 |
| V5 | R_sigma_p (ms) | 7.03 ± 5.18 | 7.27 ± 1.19 | 0.315 |
|  | R_sigma_n (ms) | 5.14 ± 1.47 | 5.77 ± 1.18 | < .0001 |
|  | T_sigma_p (ms) | 26.05 ± 6.4 | 26.19 ± 3.25 | 0.664 |
|  | T_sigma_n (ms) | 75.7 ± 20.41 | 47.64 ± 10.75 | < .0001 |
|  | RT_mean_duration_p (ms) | 308.3 ± 30.44 | 302.67 ± 27.5 | < .0001 |
|  | RT_mean_duration_n (ms) | 192.77 ± 42.03 | 216.04 ± 23.81 | < .0001 |
|  | QT_mean_duration (ms) | — | 335.36 ± 31.78 | — |
|  | R_mean_diff (ms) | 21.23 ± 4.17 | 21.23 ± 3.17 | 0.979 |
|  | RR (ms) | 917.67 ± 137.97 | 917.8 ± 138.14 | 0.375 |
|  | R_k_p | 1.41 ± 0.49 | 1.54 ± 0.5 | < .0001 |
|  | R_k_n | 1.51 ± 0.47 | 1.48 ± 0.49 | 0.000 |
|  | RMSE | 0.026 ± 0.011 | 0.006 ± 0.001 | < .0001 |

All parameters expressed as mean ± SD. *p*-values from paired *t*-tests (*df*=443). QT mean duration is unique to the 10-CDF model and has no 4-CDF counterpart; paired comparison is not applicable (—). CDF, cumulative distribution function; ms, milliseconds; RMSE, root mean square error; SD, standard deviation.

### Normative values for the 10-CDF model

Table 4 presents the normative reference values for 10-CDF TCG parameters stratified by sex for leads II, V1, and V5. Sex differences were observed for several parameters; notably, RT_mean_duration_p was significantly longer in women than in men in leads II (319.29 ± 28.35 vs. 302.81 ± 25.29 ms) and V5 (315.9 ± 28.34 vs. 298.78 ± 26.04 ms), reflecting sex-related differences in cardiac dimensions.

**Table 4.**
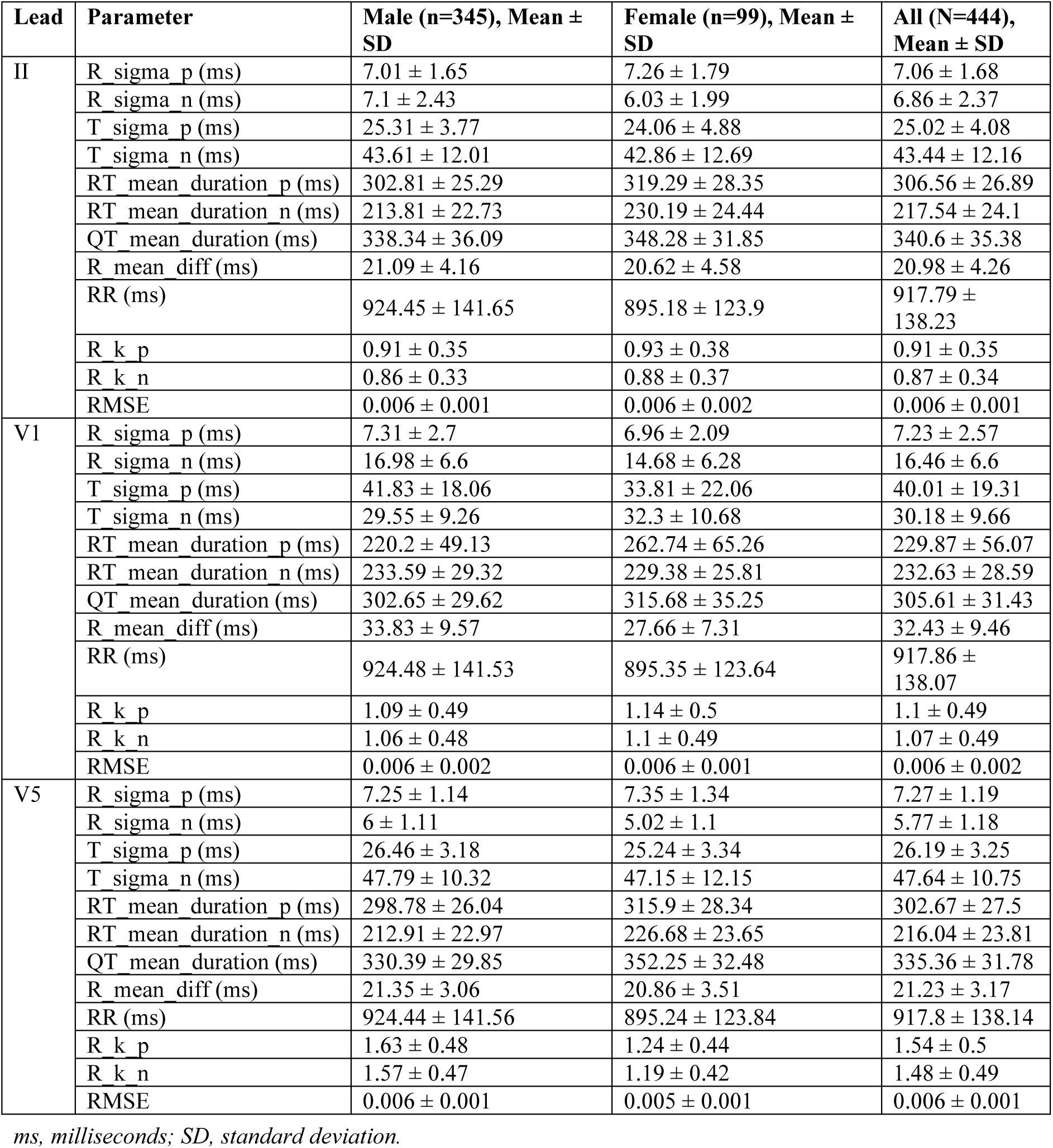
10-CDF TCG normative values (N=444)

| Lead | Parameter | Male (n=345), Mean ± SD | Female (n=99), Mean ± SD | All (N=444), Mean ± SD |
| --- | --- | --- | --- | --- |
| II | R_sigma_p (ms) | 7.01 ± 1.65 | 7.26 ± 1.79 | 7.06 ± 1.68 |
| | R_sigma_n (ms) | 7.1 ± 2.43 | 6.03 ± 1.99 | 6.86 ± 2.37 |
| | T_sigma_p (ms) | 25.31 ± 3.77 | 24.06 ± 4.88 | 25.02 ± 4.08 |
| | T_sigma_n (ms) | 43.61 ± 12.01 | 42.86 ± 12.69 | 43.44 ± 12.16 |
| | RT_mean_duration_p (ms) | 302.81 ± 25.29 | 319.29 ± 28.35 | 306.56 ± 26.89 |
| | RT_mean_duration_n (ms) | 213.81 ± 22.73 | 230.19 ± 24.44 | 217.54 ± 24.1 |
| | QT_mean_duration (ms) | 338.34 ± 36.09 | 348.28 ± 31.85 | 340.6 ± 35.38 |
| | R_mean_diff (ms) | 21.09 ± 4.16 | 20.62 ± 4.58 | 20.98 ± 4.26 |
| | RR (ms) | 924.45 ± 141.65 | 895.18 ± 123.9 | 917.79 ± 138.23 |
| | R_k_p | 0.91 ± 0.35 | 0.93 ± 0.38 | 0.91 ± 0.35 |
| | R_k_n | 0.86 ± 0.33 | 0.88 ± 0.37 | 0.87 ± 0.34 |
| | RMSE | 0.006 ± 0.001 | 0.006 ± 0.002 | 0.006 ± 0.001 |
| V1 | R_sigma_p (ms) | 7.31 ± 2.7 | 6.96 ± 2.09 | 7.23 ± 2.57 |
| | R_sigma_n (ms) | 16.98 ± 6.6 | 14.68 ± 6.28 | 16.46 ± 6.6 |
| | T_sigma_p (ms) | 41.83 ± 18.06 | 33.81 ± 22.06 | 40.01 ± 19.31 |
| | T_sigma_n (ms) | 29.55 ± 9.26 | 32.3 ± 10.68 | 30.18 ± 9.66 |
| | RT_mean_duration_p (ms) | 220.2 ± 49.13 | 262.74 ± 65.26 | 229.87 ± 56.07 |
| | RT_mean_duration_n (ms) | 233.59 ± 29.32 | 229.38 ± 25.81 | 232.63 ± 28.59 |
| | QT_mean_duration (ms) | 302.65 ± 29.62 | 315.68 ± 35.25 | 305.61 ± 31.43 |
| | R_mean_diff (ms) | 33.83 ± 9.57 | 27.66 ± 7.31 | 32.43 ± 9.46 |
| | RR (ms) | 924.48 ± 141.53 | 895.35 ± 123.64 | 917.86 ± 138.07 |
| | R_k_p | 1.09 ± 0.49 | 1.14 ± 0.5 | 1.1 ± 0.49 |
| | R_k_n | 1.06 ± 0.48 | 1.1 ± 0.49 | 1.07 ± 0.49 |
| | RMSE | 0.006 ± 0.002 | 0.006 ± 0.001 | 0.006 ± 0.002 |
| V5 | R_sigma_p (ms) | 7.25 ± 1.14 | 7.35 ± 1.34 | 7.27 ± 1.19 |
| | R_sigma_n (ms) | 6 ± 1.11 | 5.02 ± 1.1 | 5.77 ± 1.18 |
| | T_sigma_p (ms) | 26.46 ± 3.18 | 25.24 ± 3.34 | 26.19 ± 3.25 |
| | T_sigma_n (ms) | 47.79 ± 10.32 | 47.15 ± 12.15 | 47.64 ± 10.75 |
| | RT_mean_duration_p (ms) | 298.78 ± 26.04 | 315.9 ± 28.34 | 302.67 ± 27.5 |
| | RT_mean_duration_n (ms) | 212.91 ± 22.97 | 226.68 ± 23.65 | 216.04 ± 23.81 |
| | QT_mean_duration (ms) | 330.39 ± 29.85 | 352.25 ± 32.48 | 335.36 ± 31.78 |
| | R_mean_diff (ms) | 21.35 ± 3.06 | 20.86 ± 3.51 | 21.23 ± 3.17 |
| | RR (ms) | 924.44 ± 141.56 | 895.24 ± 123.84 | 917.8 ± 138.14 |
| | R_k_p | 1.63 ± 0.48 | 1.24 ± 0.44 | 1.54 ± 0.5 |
| | R_k_n | 1.57 ± 0.47 | 1.19 ± 0.42 | 1.48 ± 0.49 |
| | RMSE | 0.006 ± 0.001 | 0.005 ± 0.001 | 0.006 ± 0.001 |

ms, milliseconds; SD, standard deviation.

### Distribution characteristics of 10-CDF parameters

The distributional properties of 10-CDF parameters are presented in Table 5. Several parameters (R_sigma, T_sigma, and R_k) were better described by the log-normal distributions, whereas RT_mean_duration and T_mean_diff approximated normal distributions. Notably, RT_mean_duration_p in lead II followed a normal distribution in the 10-CDF model (Shapiro–Wilk p=0.566) but deviated significantly from normality in the 4-CDF model (p < 0.0001), suggesting better statistical properties in the 10-CDF framework.

**Table 5.** Distribution fitting results for 10-CDF parameters (Lead II,V1,V5, N=444)

| parameter | Shapiro-Wilk p |  |  |
| --- | --- | --- | --- |
|  | II | V1 | V5 |
| R_sigma_p (ms) | < .0001 | < .0001 | < .0001 |
| R_sigma_n (ms) | < .0001 | < .0001 | < .0001 |
| T_sigma_p (ms) | < .0001 | < .0001 | < .0001 |
| T_sigma_n (ms) | < .0001 | < .0001 | 0.171 |
| RT_mean_duration_p (ms) | 0.566 | < .0001 | 0.413 |
| RT_mean_duration_n (ms) | 0.081 | 0.952 | 0.323 |
| QT_mean_duration (ms) | < .0001 | 0.509 | 0.136 |
| R_mean_diff (ms) | < .0001 | < .0001 | 0.000 |
| RR (ms) | 0.050 | 0.052 | 0.049 |
| R_k_p | < .0001 | < .0001 | < .0001 |
| R_k_n | < .0001 | < .0001 | < .0001 |
| RMSE | < .0001 | < .0001 | < .0001 |

### Correlation of TCG parameters with age and body surface area

Correlation analyses (Table 6) revealed that RT_mean_duration was significantly dependent on age and body surface area (BSA). The RT_mean_duration_p increased with age (R=0.17, p=0.0002) and decreased with BSA (R=-0.21, p < 0.0001) in lead II, and showed similar patterns in lead V5 (age R=0.16, p=0.0004; BSA R=-0.21, p < 0.0001). In contrast, most T_sigma and R_sigma parameters showed nonsignificant correlations with age and BSA (|R|<0.10), indicating that these parameters reflect intrinsic myocardial electrical properties that are relatively unaffected by anthropometric factors. R_k_p and R_k_n in lead V5 showed significant positive correlations with BSA (R=0.27 and 0.28, p < 0.0001).

**Table 6.**
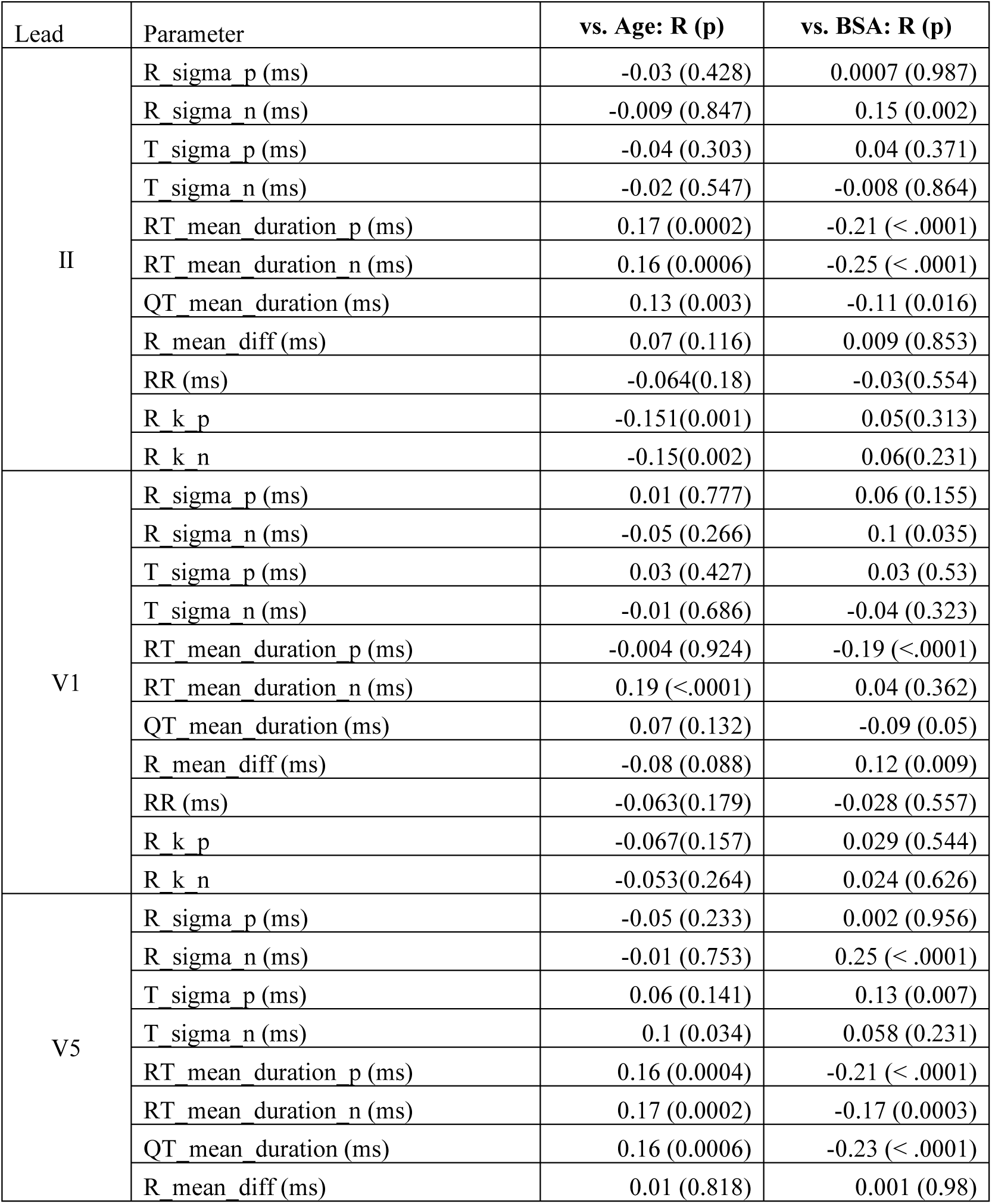
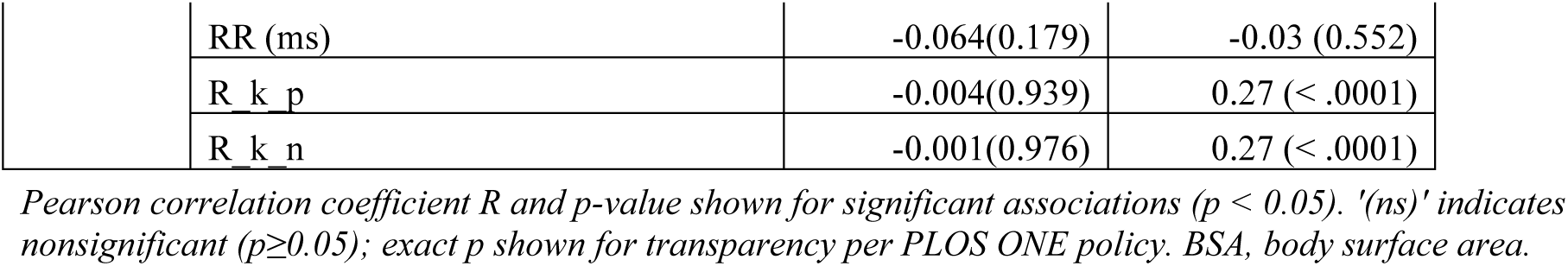
Significant correlations of 10-CDF TCG parameters with age and body surface area.

| Lead | Parameter | vs. Age: R (p) | vs. BSA: R (p) |
| --- | --- | --- | --- |
| II | R_sigma_p (ms) | -0.03 (0.428) | 0.0007 (0.987) |
|  | R_sigma_n (ms) | -0.009 (0.847) | 0.15 (0.002) |
|  | T_sigma_p (ms) | -0.04 (0.303) | 0.04 (0.371) |
|  | T_sigma_n (ms) | -0.02 (0.547) | -0.008 (0.864) |
|  | RT_mean_duration_p (ms) | 0.17 (0.0002) | -0.21 (< .0001) |
|  | RT_mean_duration_n (ms) | 0.16 (0.0006) | -0.25 (< .0001) |
|  | QT_mean_duration (ms) | 0.13 (0.003) | -0.11 (0.016) |
|  | R_mean_diff (ms) | 0.07 (0.116) | 0.009 (0.853) |
|  | RR (ms) | -0.064(0.18) | -0.03(0.554) |
|  | R_k_p | -0.151(0.001) | 0.05(0.313) |
|  | R_k_n | -0.15(0.002) | 0.06(0.231) |
| V1 | R_sigma_p (ms) | 0.01 (0.777) | 0.06 (0.155) |
|  | R_sigma_n (ms) | -0.05 (0.266) | 0.1 (0.035) |
|  | T_sigma_p (ms) | 0.03 (0.427) | 0.03 (0.53) |
|  | T_sigma_n (ms) | -0.01 (0.686) | -0.04 (0.323) |
|  | RT_mean_duration_p (ms) | -0.004 (0.924) | -0.19 (<.0001) |
|  | RT_mean_duration_n (ms) | 0.19 (<.0001) | 0.04 (0.362) |
|  | QT_mean_duration (ms) | 0.07 (0.132) | -0.09 (0.05) |
|  | R_mean_diff (ms) | -0.08 (0.088) | 0.12 (0.009) |
|  | RR (ms) | -0.063(0.179) | -0.028 (0.557) |
|  | R_k_p | -0.067(0.157) | 0.029 (0.544) |
|  | R_k_n | -0.053(0.264) | 0.024 (0.626) |
| V5 | R_sigma_p (ms) | -0.05 (0.233) | 0.002 (0.956) |
|  | R_sigma_n (ms) | -0.01 (0.753) | 0.25 (< .0001) |
|  | T_sigma_p (ms) | 0.06 (0.141) | 0.13 (0.007) |
|  | T_sigma_n (ms) | 0.1 (0.034) | 0.058 (0.231) |
|  | RT_mean_duration_p (ms) | 0.16 (0.0004) | -0.21 (< .0001) |
|  | RT_mean_duration_n (ms) | 0.17 (0.0002) | -0.17 (0.0003) |
|  | QT_mean_duration (ms) | 0.16 (0.0006) | -0.23 (< .0001) |
|  | R_mean_diff (ms) | 0.01 (0.818) | 0.001 (0.98) |

|  |  |  |
| --- | --- | --- |
| RR (ms) | -0.064(0.179) | -0.03 (0.552) |
| R<sub>k p</sub> | -0.004(0.939) | 0.27 (< .0001) |
| R<sub>k n</sub> | -0.001(0.976) | 0.27 (< .0001) |

Pearson correlation coefficient $R$ and $p$ -value shown for significant associations ( $p < 0.05$ ). '(ns)' indicates nonsignificant ( $p \geq 0.05$ ); exact $p$ shown for transparency per PLOS ONE policy. BSA, body surface area.

Notably, these correlations were observed in a deliberately narrow healthy cohort. The study eligibility criteria included a body mass index ≥ 18.5 and < 25.0 kg/m². The BMI cutoff of 18.5–25.0 kg/m² structurally excluded participants with overt obesity or marked leanness. Consequently, the resulting narrow BSA distribution (mean 1.71 m²; see Table 1) may have likely resulted in weak correlations between the TCG parameters and BSA.

### Discussion Principal findings

This study demonstrates that, in an initial evaluation conducted in healthy individuals, increasing the number of CDFs from 4 to 10 in tensor cardiography produces a highly significant improvement in ECG waveform reconstruction accuracy, with very large effect sizes (Cohen’s d=-1.84 ∼ -2.77) across representative leads. The 10-CDF model achieved RMSE values of 0.0061-0.0063 in leads II, V1, and V5, representing a 72-77% reduction compared with the 4-CDF model. The r² improved from median values of 0.952-0.971 (4-CDF) to 0.997-0.998 (10-CDF), indicating near-perfect waveform reconstruction. Beyond the RMSE improvement, the 10-CDF model yielded parameter estimates with substantially reduced variability and improved distributional properties, suggesting more stable and physiologically meaningful fitting.

### Why does increased CDF resolution improve accuracy?

This improvement can be understood from both the mathematical and physiological perspectives. Mathematically, each CDF contributes a sigmoidal function with four free parameters to the composite waveform model. With four CDFs (16 parameters per lead), the model had a limited capacity to represent the full complexity of the P-QRS-T waveform, which contained multiple overlapping components arising from distinct electrophysiological processes. With ten CDFs (40 parameters per lead), the model gained sufficient degrees of freedom to allocate dedicated CDFs to the P-wave, initial and terminal QRS components (important for notched or fragmented QRS), and late repolarization features. This is particularly relevant for the precordial leads V1-V4, where complex multiphasic QRS morphologies require more CDFs for accurate representation. The additional Q and S components in the 10-CDF framework facilitate a more accurate representation of complex QRS morphologies, thereby improving the stability and physiological interpretability of the R- and T-wave parameters.

The observation that many parameters showed reduced standard deviations in the 10-CDF model is particularly noteworthy. The 4-CDF model’s parameters appeared to absorb the fitting residuals from the unmodeled waveform components, inflating their apparent variability. The 10-CDF model, by explicitly modeling these components, produces parameter estimates that more precisely reflect the true inter-individual physiological variation.

### Clinical significance

The establishment of normative reference values in 444 healthy individuals represents a foundational step toward clinical application. These values define the ‘normal tensor space’ against which pathological deviations can be measured. Prior work has shown that conventional automated ECG algorithms have inherent limitations in sensitivity to subtle abnormalities [3,15]; the 10-CDF TCG framework may enable more sensitive detection of early pathological changes.

The identification of the two new parameters, T_mean_diff and RT_mean_duration, was found to be clinically significant. T_mean_diff provides a novel index of repolarization asymmetry and RT_mean_duration quantifies the temporal span of the depolarization-to-repolarization process more precisely than the conventional QT interval. TCG has already demonstrated utility in detecting myocardial ischemia and HCM using the 4-CDF framework [14]; the 10-CDF model may further enhance these applications by reducing fitting residual ‘noise’ and providing additional discriminating parameters. Furthermore, we are currently developing a high-precision classification model based on LightGBM, in which features derived from 10CDF-TCG are used to identify specific cardiac conditions, including myocardial ischemia, hypertrophic cardiomyopathy, and cardiac sarcoidosis. By leveraging quantitative TCG metrics, this approach is expected to facilitate explainable AI-based analysis and enhance the interpretability of ECG-derived diagnostic outputs. TCG also functions as a data compression method: a continuous ECG waveform (≈500-1000 data points per lead per beat) is reduced to a compact physiologically interpretable parameter set. A 4-CDF TCG compression rate of approximately 9% (data size 9/100) was previously shown to be superior to the standard DCT at a comparable reconstruction quality [14]; the 10-CDF model retains a comparable compression efficiency while substantially improving reconstruction fidelity, making the 10-CDF model suitable for long-term ECG storage, personal health records, and real-time remote monitoring systems.

The correlation analyses revealed that while RT_mean_duration shows significant age and BSA dependence—requiring appropriate normalization in clinical application—most σ and κ parameters are relatively independent of these confounders, making them potentially robust biomarkers independent of body habitus.

### Comparison with existing methods

Transform-based ECG compression methods typically achieve compression ratios of 8:1 to 20:1 with percent root-mean-square difference values of 0.25-6% [6,7,16]. The 10-CDF TCG model, with typical RMSE values of 0.006-0.007 and r²>0.996, achieves a reconstruction accuracy comparable to or superior to these methods, while providing a physiologically structured representation entirely absent from transform-based approaches. Direct systematic benchmarking against established ECG compression standards is required in future studies.

### Demographic moderation of TCG parameters and impact of the healthy-cohort design

Conventional electrocardiographic parameters are substantially modulated by age and body size. Multiple large cohort studies have shown that the QTc interval increases with advancing age, with a steeper relationship in men than in women, and that healthy reference ranges differ markedly according to age and sex [17,18]. T-wave amplitude declines with age—approximately 10% between 18 and 39 and 40–59 decades and an additional 15% between 40 and 59 and 60–79 years —and is, on average, 25% larger in men, reflecting greater cardiac mass and repolarizing-ion-channel expression [19]. Body habitus likewise affects precordial

QRS amplitudes, primarily through the electrically insulating effect of thoracic adipose tissue. Sokolow–Lyon voltage is attenuated in obesity, whereas Cornell-voltage and voltage–duration product criteria are relatively preserved or even increased [20,21]. Obesity is also associated with mild QT/QTc prolongation through heightened sympathetic activation and reduced heart-rate variability [21]. More recently, 3D QRS voltage–time integrals have been shown to depend strongly on age and sex in healthy adults, prompting age- and sex-specific reference ranges, whereas their association with BSA is comparatively weak [22].

Against this background, our finding that most TCG shape parameters (T_sigma and R_sigma; |R|<0.10) showed no meaningful correlation with age or BSA, while only the duration-type parameters (RT_mean_duration, QT_mean_duration) and selected V5 amplitude-shape parameters (R_k_p, R_k_n) showed weak to modest correlations (|R| up to ∼0.27) should be interpreted in the context of our deliberately narrow inclusion criteria. Consequently, individuals with overt obesity, marked leanness, hypertensive remodeling, or subclinical left ventricular hypertrophy are largely absent. In particular, the BMI cutoff of 18.5–25.0 kg/m² structurally excluded outliers at both ends of the body-size spectrum, and the resulting narrow distributional range of BSA (mean 1.71 m²) likely attenuated the correlation coefficients observed here, and the present analysis should therefore be regarded as establishing the baseline behavior of 10-CDF TCG parameters in unequivocally healthy individuals rather than as a comprehensive characterization of their full demographic dependence. The two implications are as follows. First, the relative independence of the T_sigma and R_sigma shape parameters from anthropometric factors, even with restricted sampling, suggests that these higher-order shape descriptors may primarily capture intrinsic myocardial electrical properties rather than torso-related transmission effects, which is attractive for cross-population diagnostic comparisons. Second, the duration-type parameters (RT_mean_duration and QT_mean_duration), which already showed age- and BSA-modulation in this homogeneous cohort, will likely require explicit age-, sex-, and possibly BSA-specific normative ranges when extended to broader populations, analogous to the existing recommendations for QTc and QRS-3D voltage indices [18,22]. A planned follow-up study in a demographically diverse cohort, including overweight, obese, and underweight participants, is required to define such ranges and test whether the apparent anthropometric independence of T_sigma and R_sigma is preserved beyond a narrowly screened healthy population.

### Integration with deep-learning ECG analysis

Deep learning has driven rapid progress in ECG analysis, with one-dimensional convolutional architectures such as ResNet and Inception achieving high PTB-XL performance [8]. Yet these models demand substantial compute and offer limited explainability — barriers to clinical adoption, since physicians require an understandable rationale alongside accurate prediction. Signal representation strongly shapes both performance and cost, motivating diverse front-end approaches including time-, frequency-, time–frequency-, decomposition-, sparse-, deep-feature-, image-, wavelet/scalogram-, and tensor-based methods [9].

An ideal ECG representation should compress the waveform without losing diagnostically critical content, permit morphological reconstruction, and yield physiologically meaningful features — the principle underlying explainable-by-design approaches such as FactorECG [10]. By contrast, post-hoc saliency or heatmap methods can be unstable, may not faithfully reflect a model’s decisions, and are judged by clinicians as only modestly informative [11].

TCG addresses these requirements directly: it models collective myocardial action-potential transitions as differences between endocardial and epicardial cumulative distribution functions, organized as a fourth-order tensor of means, variances, weights, and levels across beats and leads [14]. The fitted components reconstruct waveform segments, enabling direct comparison with morphological abnormalities. When paired with tree-based gradient boosting (XGBoost, LightGBM) and SHAP, TCG features yield both efficiency and interpretability; a recent benchmark study reported a macro AUC of 0.933 — close to ResNet (0.937) and above LSTM (0.927) — with explicit feature-importance attribution by lead, wave component, and parameter type [12].

Such lightweight, interpretable processing fits time-critical settings (e.g., the 10-minute 12-lead ECG target in suspected acute coronary syndrome) and embedded, ambulatory, and consumer-grade devices (Holter, patch, smartphone- and smartwatch-linked) now covered by Japanese consensus guidance [13]. Edge-side processing is increasingly required where latency, energy, memory, or privacy constrain cloud inference. TCG therefore offers a promising low-resource, physiologically grounded representation, pending broader external validation and benchmarking against transformer and foundation-model approaches.

### Limitations

This study had some limitations. First, this study exclusively analyzed ECGs from rigorously screened healthy individuals; the performance in the presence of pathological waveforms remains to be established. Second, the population in the occupational screening cohort was predominantly male (77.7 %) and of Japanese ethnicity, limiting generalizability. Third, the 60–79 age group was underrepresented (n=39). Fourth, a formal paired t-test analysis with Cohen’s d was performed for leads II, V1, and V5 as representative, and an extension to all 12 leads is warranted. Fifth, a detailed 10-CDF decomposition scheme, including the CDF allocation to specific waveform components, requires full specifications for reproducibility. Our understanding of P-wave analysis, its extension to more complex waveforms, and its relationship with U-waves remains incomplete and may be improved by increasing the number of CDF components. Finally, the principal aim of the present study was to evaluate the reconstruction accuracy of the 10-CDF tensor cardiography model in rigorously screened healthy individuals and to establish baseline normative parameter ranges; systematic assessment of sex differences and of the influence of obesity and other anthropometric or metabolic factors on 10-CDF TCG parameters was therefore beyond the scope of this work and is planned as a separate investigation in a demographically and metabolically more diverse cohort.

### Conclusions

Increasing the CDF resolution from 4 to 10 in TCG significantly enhanced the accuracy and stability of ECG waveform reconstruction in healthy individuals, achieving median r² values exceeding 0.996 and RMSE values of 0.006-0.007 across representative leads—a 72-77% improvement over the 4-CDF model with very large effect sizes (Cohen’s d=-1.84 ∼ -2.77). The 10-CDF model produces parameter estimates with reduced variability and improved distributional properties, and enables the extraction of novel parameters (T_mean_diff, RT_mean_duration) that are not available in the 4-CDF framework. The normative reference values established in this study for 444 healthy Japanese adults provide a foundation for future clinical validation in diseased populations.

## Materials and methods

### Study population

This study analyzed standard 12-lead ECGs for 444 healthy individuals (345 men and 99 women; age range 20–79 years) obtained from two sources: (1) Tobu Railway Co. Ltd. Corporation occupational health screening via the Katsushika Health Screening Center (from approximately 3,700 employees screened, 415 met all inclusion criteria), and (2) Nippon Medical School Hospital ECG database (from 1,958 cases, 29 individuals met all inclusion criteria). Participants with known cardiac diseases, electrocardiogram (ECG) abnormalities, or medications affecting ECG morphology were excluded from the study.

The following eligibility criteria were applied identically to both source cohorts: 1) Medical history: no coronary artery disease, arrhythmia, or other cardiac disease; 2) Lifestyle: Non-smoking; 3) Anthropometric and laboratory thresholds at annual health screening: body mass index 18.5 to < 25.0 kg/m²; systolic blood pressure 90–130 mmHg and diastolic blood pressure ≤ 84 mmHg; fasting plasma glucose 60–109 mg/dL; HbA1c ≤ 5.8%; serum uric acid ≤7.0 mg/dL; and estimated glomerular filtration rate ≥ 60 mL/min/1.73 m²; and 4) ECG morphology classified as normal by an automated diagnostic software (Fukuda Denshi) and readjudicated as normal by board-certified cardiologists (Y.M. and C.N. for waveform confirmation, and K.Y., Y.I., and Y.T.T. for data extraction and clinical review). These criteria were applied in a stepwise fashion for the Tobu Railway cohort: of approximately 3,700 examinees, 940 satisfied all blood test criteria (overall grade A). A case-by-case ECG review further narrowed this set to 525 participants. Additionally, exclusion of hypertension, cardiac disease, and other chronic diseases finally yielded 415 individuals for analysis. These thresholds structurally excluded participants with overt obesity, marked leanness, hypertensive or prehypertensive states, dysglycemia, hyperuricemia, or reduced renal function. The narrow eligibility window was selected to establish normative 10-CDF TCG reference values in unequivocally healthy individuals while acknowledging that it constrains the dynamic range of anthropometric and metabolic covariates available for correlation analyses.

The study was conducted in accordance with the Declaration of Helsinki and the Belmont Report. The study protocol was approved by the Central Ethics Committee of Nippon Medical School (Approval No. M-2023-159). This study was conducted as part of an Agency for Medical Research and Development (AMED)-funded project on the high-precision early automatic diagnosis of hereditary cardiac diseases using tensor electrocardiography. The requirement for individual consent was waived by the ethics committee because of the retrospective nature of the analysis.

### ECG acquisition and data processing

All participants underwent electrocardiography measurements using standard 12-lead ECGs recorded at rest using Fukuda Denshi electrocardiographs following standard lead placement protocols [23]. All recordings were performed in the supine position after at least 5 min of rest. Raw ECG data were converted to a digital format at a sampling rate of 500 Hz, suitable for TCG analysis by the Fukuda Denshi Corporation.

### Tensor cardiography analysis

#### Theoretical framework

TCG represents the ECG waveform as a linear combination of CDFs based on the physiological principle that collective transitions of ventricular myocardial action potentials approximate Gaussian distributions [14]. The depolarization process (R wave) is modeled as follows:

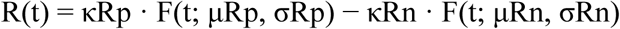

where F(t; μ, σ) denotes the CDF of a Gaussian distribution with mean μ and standard deviation σ, and κ represents the weight of each component. Similarly, the T-wave is represented by the inverse CDFs for the repolarization process. The ECG dipole model and the R- and T-wave parameters derived from this framework are illustrated in Fig 2.

**Fig.2.**
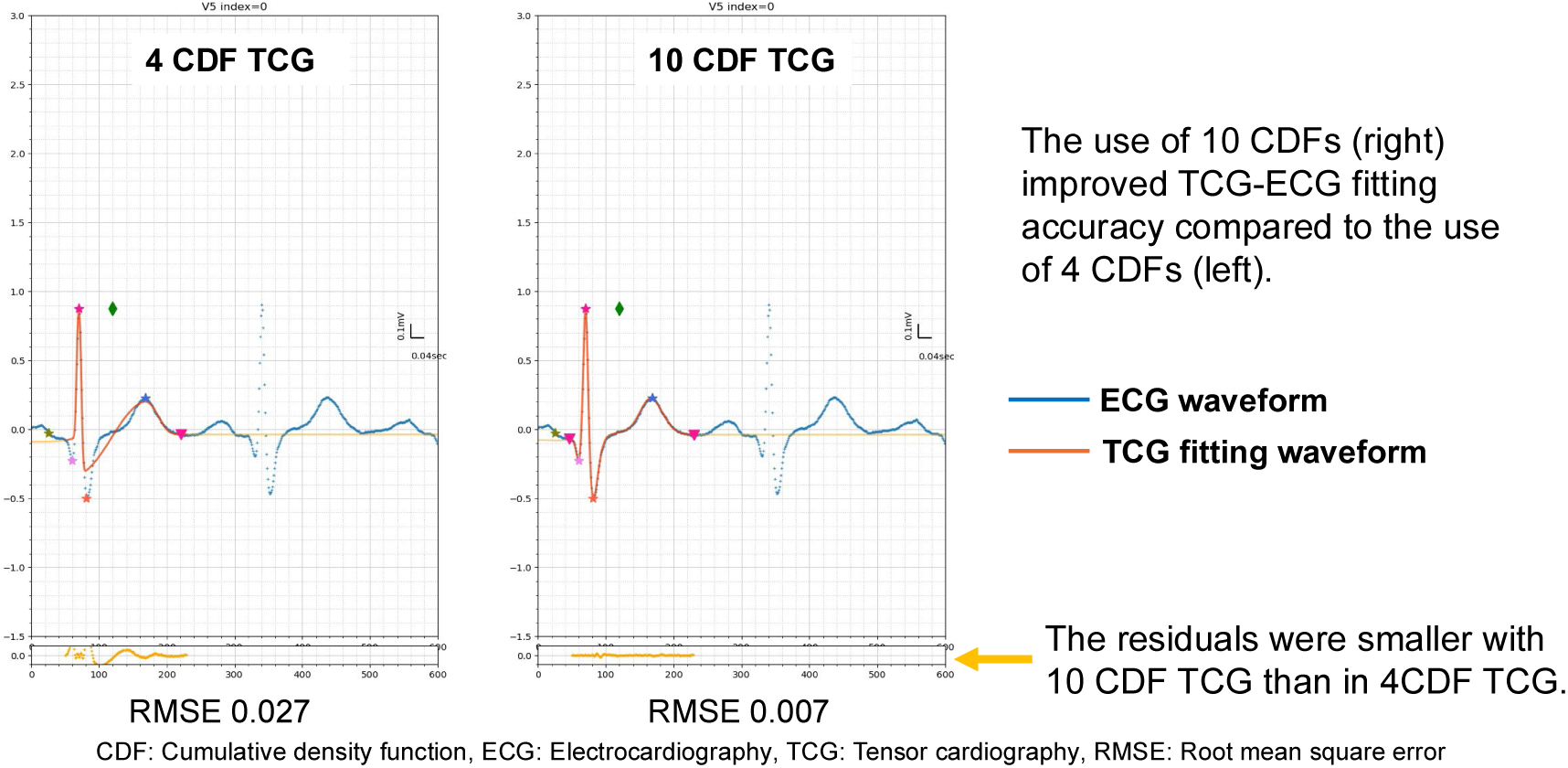
Representative ECG waveform reconstructions using the 4-CDF and 10-CDF TCG models. Upper panels: original ECG waveform (blue), 4-CDF reconstruction (left), and 10-CDF reconstruction (right). Lower panels: residual traces (yellow). The markedly smaller residuals with the 10-CDF model are evident across the P-QRS-T complex. CDF, cumulative distribution function; ECG, electrocardiogram; TCG, tensor cardiography.

**Fig 2.**
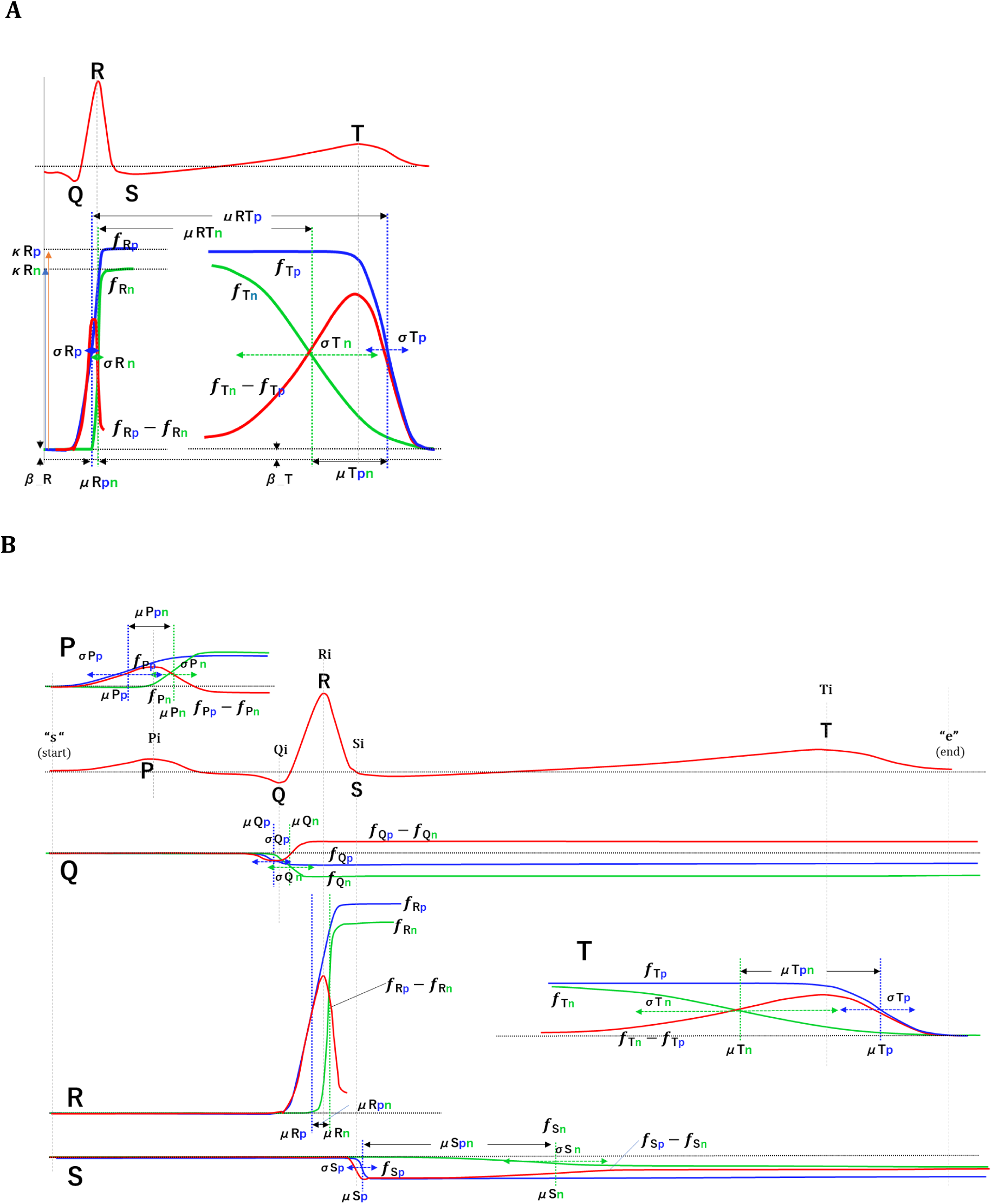
**A.** Detailed parameter definitions based on the 4-CDF model illustrated on the fitted CDF waveforms for R- and T-wave regions: σ (spread) and μ (mean timing) for the positive and negative components of the R wave (σRp, σRn, μRp) and T wave (σTp, σTn, μTp, μTn); κ (relative amplitude / weight, κR on the R-wave panel); and β (baseline level, βR and βT). μRT denotes the R-to-T mean-timing interval (A). In this study, software notation is used: σRp=R_sigma_p, σRn=R_sigma_n, σTp=T_sigma_p, σTn=T_sigma_n, μTp−μRp=RT_mean_duration_p, μTn−μRn=RT_mean_duration_n, μRp−μRn=R_mean_diff, μTp−μTn=T_mean_diff. Adapted from Tsukada et al. [14]. **B.** Parameter definitions based on the 10-CDF model. The six additional CDF sets—three positive and three negative—were added to the 10-CDF model to represent the P, Q, and S waves. CDF, cumulative distribution function; ECG, electrocardiogram; TCG, tensor cardiography.

### Tensor formulation

The TCG framework constitutes a fourth-order tensor T ∈ ℝ^L^ ^×^ ^C^ ^×^ ^P^ ^×^ ^N^, where the four modes index: (1) ECG lead l ∈ {I, II, III, aVR, aVL, aVF, V1–V6}, (2) CDF component c ∈ {1, …, 10}, (3) waveform parameter p ∈ {μ, κ, σ, β} (mean timing, weight, spread, baseline level), and (4) heartbeat index n ∈ {1, …, N}. For the current cohort, the dimensions were 12 × 10 × 4 × N (12 leads × 10 CDFs × 4 parameters × N beats), and each entry T(l, c, p, n) denotes the value of parameter p for the cth CDF component of lead l in the nth beat. This tensor representation, from which the term *Tensor Cardiography* is derived, enables joint analysis across leads, CDF components, parameters, and time without collapsing any mode prematurely, and provides a natural data structure for downstream multilinear and machine-learning analyses.

#### The 4-CDF model

In the standard 4-CDF model, the ECG is decomposed into four CDFs: fRp and fRn for the R wave (depolarization), and f’Tp and f’Tn for the T wave (repolarization). Each CDF yields four parameters (σ, μ, κ, β), producing 16 parameters per lead per beat.

#### The 10-CDF model

The 10-CDF model extends the standard framework by incorporating six additional CDFs to capture waveform components that are not explicitly modeled in the 4-CDF configuration, including the P wave, finer QRS morphological features, and late repolarization components (Fig 2B). The 10-CDF model enables the extraction of two additional derived parameters that are not available in the 4-CDF framework: T_mean_diff (the mean difference index for the T-wave, characterizing repolarization asymmetry) and RT_mean_duration_p/n (the mean duration index for the positive and negative components of the depolarization-to-repolarization interval).

#### Fitting procedure

TCG fitting was performed for each participant using the 4-CDF and 10-CDF models in leads II, V1, and V5 among the 12 leads using the TCG analysis software. The least squares method was used to minimize the residual difference between the observed and model-reconstructed ECG waveforms. The median beat from the R-R interval series was selected as the representative beat for the analysis [14].

### Evaluation metrics

#### Reconstruction accuracy

The primary endpoint was the RMSE between the original ECG waveform and the TCG-reconstructed waveform normalized to the waveform amplitude scale (mV). The coefficient of determination (r²) was calculated to express the proportion of the waveform variance explained by the TCG model.

#### TCG parameters

The following parameters were extracted per lead (Table 7; Fig 2): standard deviation (σ) of the R-wave positive and negative components (R_sigma_p, R_sigma_n) and T-wave positive and negative components (T_sigma_p, T_sigma_n); RT mean duration for positive and negative components (RT_mean_duration_p, RT_mean_duration_n); R-wave mean amplitude difference (R_mean_diff); T-wave mean amplitude difference (T_mean_diff); RR interval (RR); and R-wave curvature for positive and negative components (R_k_p, R_k_n).

**Table 7.** TCG parameter definitions extracted in the 10-CDF model.

| Parameter | Definition |
| --- | --- |
| R_sigma_p ($\sigma_{Rp}$) | Spread of the positive R-wave CDF component (index of depolarization synchrony, endocardial side) |
| R_sigma_n ($\sigma_{Rn}$) | Spread of the negative R-wave CDF component (epicardial side) |
| T_sigma_p ($\sigma_{Tp}$) | Spread of the positive T-wave CDF component (duration of endocardial repolarization) |
| T_sigma_n ($\sigma_{Tn}$) | Spread of the negative T-wave CDF component (epicardial repolarization) |
| RT_mean_duration_p ($\mu_{Tp}-\mu_{Rp}$) | Endocardial activation–repolarization time (mean timing from R-wave to T-wave positive component) |
| RT_mean_duration_n ($\mu_{Tn}-\mu_{Rn}$) | Epicardial activation–repolarization time (mean timing from R-wave to T-wave negative component) |
| R_mean_diff ($\mu_{Rp}-\mu_{Rn}$) | Difference in depolarization timing between endocardium and epicardium (R-wave upstroke–downstroke interval) |
| T_mean_diff ($\mu_{Tp}-\mu_{Tn}$) | Difference in repolarization timing between endocardium and epicardium (T-wave upstroke–downstroke interval) |
| R_k_p ($k_{Rp}$) | Relative amplitude contribution of the positive R-wave CDF component to QRS amplitude |
| R_k_n ($k_{Rn}$) | Relative amplitude contribution of the negative R-wave CDF component to QRS amplitude |
| RR | RR interval (beat-to-beat cycle length, ms) |

*Parameters are listed in software notation with corresponding Greek symbol notation in parentheses. All timing parameters are in milliseconds (ms) unless stated otherwise. CDF, cumulative distribution function; TCG, tensor cardiography.*

### Statistical analysis

Continuous variables are expressed as mean ± standard deviation (SD). Normative ranges were defined as mean ± 3SD. Data distribution characteristics were assessed using the Shapiro–Wilk test for normality and the Anderson–Darling test for normal and log-normal distributions, with the Akaike information criterion (corrected) used for model comparison. Comparisons between the 4-CDF and 10-CDF models were performed using paired t-tests, because each participant’s ECG was analyzed using both methods, yielding naturally paired observations. Cohen’s d effect sizes were calculated as the mean paired difference divided by the standard deviation of the paired differences to quantify the magnitude of the differences (|d|>0.8 classified as a large effect). Pearson’s correlation coefficients were used to assess the relationships between TCG parameters and age or body surface area (BSA). Statistical significance was defined as a two-sided p < 0.05. All analyses were performed using JMP Pro software (SAS Institute, Cary, NC, USA). Statistical significance was set at p =0.05.

## Data Availability

For Study Protocols: No datasets were generated or analysed during the current study. All relevant data from this study will be made available upon study completion.

## Acknowledgments

The authors thank Tobu Railway Co., Ltd. for their cooperation in providing occupational health screening data, Takao Katoh (Tobu Railway Health Screening Center) for coordinating health screening participants, and the Katsushika Health Screening Center for participant coordination. The authors would also like to express their gratitude to Yuko Miyazoe and Chiaki Nakamura for their assistance with the data cleaning and analysis.

## Author contributions

Conceptualization: S. Tsukada, Y.T. Tsukada, and Y. Iwasaki. Data curation: K. Yodogawa, Y. Iwasaki and Y.T. Tsukada. Formal analysis: Tsukada S and Shiozawa A Funding acquisition: Y.T. and Tsukada. Investigation: H. Hirayama, K. Yodogawa, H. Murata, Y. Iwasaki and T. Fujino. Methodology: S. Tsukada and A. Shiozawa. Project administration: Y. T. Tsukada and Y. Iwasaki. Tsukada S and Shiozawa A Supervision: Y. T. Tsukada, Y. Iwasaki, and S. Tsukada. Validation: Murata and Hirayama. Visualization: H. Hirayama and S. Tsukada. Writing the original draft: Y. T. Tsukada. Writing, review, and editing: Y. T. Tsukada, Y. Iwasaki, T. Fujino, and H. Murata.

## Data availability

The raw ECG data supporting the findings of this study cannot be shared publicly because of ethical restrictions and data privacy regulations governing occupational health screening data. The aggregated statistical data presented in this manuscript are available in the article and the supplementary material. Data access requests are directed to the corresponding author.

---

**S1 Table.** Complete basic statistics for all 10-CDF TCG parameters across all 12 leads (mean, SD, SEM, 95% CI, mean±3SD, quartiles, min, max). N=444.

**S2 Table.** Complete basic statistics for all 4-CDF TCG parameters across all 12 leads. N=444.

**S3 Table.** Complete paired t-test results comparing the 4-CDF and 10-CDF TCG parameters across all analyzed leads.

**S4 Checklist.** STROBE checklist for observational study reporting.

**S1 Fig.** Distribution histograms with sex stratification (all, male and female) for all 10-CDF parameters across leads II, V1, and V5. Normal and log-normal fit curves are shown.

**S2 Fig.** Complete scatter plots of all 10-CDF TCG parameters versus age and BSA for leads II, V1, and V5.

## References

1. Kligfield P, Gettes LS, Bailey JJ, Childers R, Deal BJ, Hancock EW, et al. Recommendations for the standardization and interpretation of the electrocardiogram. Part I: The electrocardiogram and its technology. Circulation. 2007;115:1306–1324.

2. Rautaharju PM, Surawicz B, Gettes LS, Bailey JJ, Childers R, Deal BJ, et al. AHA/ACCF/HRS recommendations for the standardization and interpretation of the electrocardiogram. Part IV: The ST segment, T and U waves, and the QT interval. Circulation. 2009;119:e241–e250.

3. Schläpfer J, Wellens HJ. Computer-interpreted electrocardiograms: benefits and limitations. J Am Coll Cardiol. 2017;70:1183–1192.

4. Yodogawa K, Morita N, Kobayashi Y, Takayama H, Ohara T, Seino Y, et al. High-frequency potentials developed in wavelet-transformed electrocardiogram as a novel indicator for detecting Brugada syndrome. Heart Rhythm. 2006;3:1436–1444.

5. Takayama H, Yodogawa K, Katoh T, Takano T. Evaluation of arrhythmogenic substrate in patients with hypertrophic cardiomyopathy using wavelet transform analysis. Circ J. 2006;70:69–74.

6. Batista LV, Melcher EUK, Carvalho LC. Compression of ECG signals by optimized quantization of discrete cosine transform coefficients. Med Eng Phys. 2001;23:127–134.

7. Ahmed SM. ECG signal compression using combined modified discrete-cosine and discrete-wavelet transforms. J Eng Sci Assiut Univ. 2005;33:2111–2122.

8. Strodthoff N, Wagner P, Schaeffter T, Samek W. Deep learning for ECG analysis: benchmarks and insights from PTB-XL. IEEE J Biomed Health Inform. 2021;25(5):1519–1528.

9. Berkaya SK, Uysal AK, Gunal ES, Ergin S, Gunal S, Gulmezoglu MB. A survey on ECG analysis. Biomed Signal Process Control. 2018;43:216–235.

10. van de Leur RR, Bos MN, Taha K, Sammani A, Yeung MW, van Duijvenboden S, et al. Improving explainability of deep neural network-based electrocardiogram interpretation using variational auto-encoders. Eur Heart J Digit Health. 2022;3(3):390–404.

11. Adebayo J, Gilmer J, Muelly M, Goodfellow I, Hardt M, Kim B. Sanity checks for saliency maps. Adv Neural Inf Process Syst. 2018;31:9505–9515.

12. Saisho O, Shiozawa A, Tsukada S, Ohba T. Lightweight estimation of myocardial infarction using tensor cardiography analysis and machine learning. Proc Annu Conf JSAI. 2025;39:3H4OS10b02. doi:10.11517/pjsai.JSAI2025.0_3H4OS10b02

13. Ikeda T, Ashihara T, Iwasaki Y, et al. 2025 JHRS/JCS consensus statement on appropriate use of portable/wearable electrocardiographs. J Jpn Soc Electrocardiol. 2024;44(4):275–307. doi:10.5105/jse.44.275

14. Tsukada S, Iwasaki Y, Tsukada YT. Tensor cardiography: a novel ECG analysis of deviations in collective myocardial action potential transitions based on point processes and cumulative distribution functions. PLOS Digit Health. 2024;3:e0000273.

15. Guglin ME, Thatai D. Common errors in computer electrocardiogram interpretation. Int J Cardiol. 2006;106:232–237.

16. Jalaleddine SMS, Hutchens CG, Strattan RD, Coberly WA. ECG data compression techniques—a unified approach. IEEE Trans Biomed Eng. 1990;37:329–343.

17. Rabkin SW, Cheng XBJ, Thompson DJS. Detailed analysis of the impact of age on the QT interval. J Geriatr Cardiol. 2016;13:740–748.

18. Mason JW, Ramseth DJ, Chanter DO, Moon TE, Goodman DB, Mendzelevski B. Electrocardiographic reference ranges derived from 79,743 ambulatory subjects. J Electrocardiol. 2007;40:228–234.

19. Macfarlane PW, Lawrie TDV. T wave amplitudes in normal populations: variation with ECG lead, sex, and age. J Electrocardiol. 1995;28:191–197.

20. Okin PM, Roman MJ, Devereux RB, Pickering TG, Borer JS, Kligfield P. Time-voltage QRS area of the 12-lead electrocardiogram: detection of left ventricular hypertrophy. Hypertension. 2000;35:36–43.

21. Simonyi G. Electrocardiological features in obesity: the benefits of body surface potential mapping. Cardiorenal Med. 2014;4:123–129.

22. Beela AS, Tichy A, Stark T, Hassani-Ardakany H, Walz J, Iyer V, et al. QRS 3D voltage-time integral in narrow QRS complex: a marker for structural heart disease. J Electrocardiol. 2025;90:153897.

23. International Electrotechnical Commission. IEC 60601-2-25:2011. Medical electrical equipment – Part 2-25: Particular requirements for the basic safety and essential performance of electrocardiographs. Geneva: IEC; 2011.

